# Daily hospital outpatient visits prediction based on Seasonal Autoregressive Integrated Moving Average model

**DOI:** 10.1101/2025.04.15.25325872

**Authors:** Longting Cao, Li Zhang

## Abstract

**Objectives:** This study aimed to create a straightforward and rapid tool for predicting daily outpatient visits across the entire hospital, capturing the inherent fluctuation patterns of outpatient visits to offer a reference for the overall operation of hospital outpatient services.

**Method:** The daily outpatient visits recorded by the information system of a tertiary grade A general hospital from January 1, 2023 to December 31, 2024 was collected. Using data from January 1, 2023 to November 30, 2024, a Seasonal Autoregressive Integrated Moving Average (SARIMA) model was established to predict the daily outpatient visits in December 2024.

**Results:** The number of outpatients showed obvious cyclical characteristics and weekly seasonal trends. On the whole, the number of outpatient visits on Monday and Saturday is significantly higher than that on the other five days of the week, the number of outpatient visits on Sunday is the lowest, while the number of outpatient visits on Wednesday is slightly higher than that on Thursday and Friday. The SARIMA(1,1,0)(0,1,1)_7_ was the optimal model with strong fitting capabilities and good predictive performance, the mean absolute percentage error (MAPE) of the predicted result was 0.073.

**Conclusions:** The SARIMA model can effectively simulate the trend of changes in hospital outpatient numbers over time series, providing a theoretical basis for short-term forecasting of hospital outpatient visits. This can not only help hospitals optimize internal resource allocation and improve operational efficiency, but also provide patients with a better and more efficient medical experience.

**WHAT IS ALREADY KNOWN ON THIS TOPIC:** ⇒ Medium and long term forecasts cannot offer adequate support for the precise allocation of medical resources at the operational level. Accurate short-term forecasts can offer theoretical support for managers to schedule human resources and prepare for emergencies, which is often more practical.

**WHAT THIS STUDY ADDS:** ⇒ Our study indicates that a rapid tool for predicting daily outpatient visits can be developed by incorporating the weekly seasonal effect on outpatient numbers, while also capturing the inherent fluctuation patterns.

**HOW THIS STUDY MIGHT AFFECT RESEARCH,PRACTICE OR POLICY:** ⇒ This study can help hospital managers arrange the working hours of medical staff reasonably, and deploy medical supplies such as drugs, medical equipment, etc. to ensure patients’ medical needs.

⇒ Guide chronic disease patients to seek medical attention during off peak hours, alleviate the pressure of high outpatient visits during peak periods, and shorten patients’ waiting time.

## INTRODUCTION

The healthcare sector is of paramount importance in any society, in the present limited medical resources, efficient hospital management is very important to improve the quality of medical services.^1^ The number of outpatient visits in a hospital is the main indicator for measuring the development of a hospital’s outpatient business and an important reference for formulating medium and long-term development plans for the hospital. As a key index reflecting the state of hospital operation, accurately predicting the number of outpatient visits is crucial for efficient hospital operations. Accurately grasp the changing trend of outpatient visits, hospital managers can arrange the working hours of medical staff reasonably, and avoid the situation of manpower shortage or redundancy. It can also scientifically deploy medical supplies such as drugs, medical equipment, etc. to ensure patients’ medical needs.^2^

China has a vast population, and the rapid growth of aging demographics has made the treatment of common and chronic diseases an increasing challenge for its healthcare system. However, due to relatively scarce and unevenly distributed medical resources, accurately forecasting future healthcare demand has become crucial. With the further improvement of hospital management refinement, statistical prediction has gradually become an important means and technical support for modern hospital management work.^3 4^ Seo, et al. developed a multi-level regression model (two-level regression model) was developed for the prediction of conjunctivitis outpatient rate.^5^ Luo et al. formulated an Autoregressive Integrated Moving Average (ARIMA) model combined with a single exponential smoothing (SES) model on the day of the week time series to forecast hospital daily outpatient visits in two internal medicine departments.^6^ The Long Short-Term Memory (LSTM) and Gated Recurrent Unit (GRU) models for conjunctivitis outpatient visits prediction were established by wang et al.^7^ However, in the past few years, although there has been a growing focus on using time series models to predict the demand for medical services, most of these models are established based on weekly or monthly data.^2 8^ Alternatively, scholars concentrate on a specific department, such as the internal medicine,^9^ nursing care.^10^ Therefore, this study aims to create a straightforward and rapid tool for predicting daily outpatient volumes across the entire hospital, capturing the inherent fluctuation patterns of outpatient visits to offer a reference for the overall operation of hospital outpatient services.

Hospital outpatient visits shows strong periodical, seasonal and stochastic fluctuations driven by factors such as personal experience, various environmental factors, and the type of the sociodemographic effects.^11^ In terms of model selection, mining the key time series features of data is the premise of establishing accurate prediction models. The ARIMA model is a powerful tool in time series analysis, which enables us to capture the patterns and trends in the historical data of outpatient visits. The ARIMA has a significant advantage in processing data with time sequence. Through deep mining of historical data, it can effectively capture the characteristics of trend, seasonality and periodicity in the data.^12^ Consequently, this model has gained extensive application in forecasting research.^13^ The ARIMA model have been used in previous studies to predict monthly hospital outpatient visits based on meteorological environmental factors.^14^ Chen et al. utilized ARIMA to achieve long-term projections of emergency department revenue and visitor numbers.^15^ Considering the advantages discussed above, the ARIMA model can also be utilized for accurate short-term prediction of daily outpatient visits, providing theoretical support for managers to make rational resource allocations.

The study took place in an influential tertiary grade A general hospital in China. In this study, the ARIMA model was employed to fit the daily outpatient visits of the hospital, and an optimal predictive model was developed to estimate the daily outpatient visits. The forecasting outcomes provide a scientific foundation for hospitals to rationally allocate outpatient resources across different cycles, thereby enhancing the quality and efficiency of outpatient management.

## METHODS

### Data collection

We collected outpatient visits data from the tertiary grade A general hospital from January 1, 2023 to December 31, 2024 as our study subjects. The dataset was composed of the daily number of outpatient visits. These data were aggregated as secondary data without any personal information and therefore informed consent was not required. There were no missing values in this dataset.

### Statistical analysis

After data collection and preprocessing, the ARIMA model was applied to the modeling of this dataset. ARIMA model was composed of autoregression (AR) with a lag number denoted by p, integrate (I) with a lag number denoted by d, and moving average (MA) with a lag number denoted by q. AR indicates that current observations are correlated with previous ones, which provides a possibility of predicting with a time trend. MA refers to the correlation between the errors as well as the weighted average of random disturbance terms.^16^ When the time series presents seasonal trend, the multiplicative seasonal autoregressive comprehensive moving average model SARIMA(p, d, q)(p, d, q) s can be used.^17^ The seasonal autoregressive integrated moving average (SARIMA) model is an extension of the ARIMA model, which includes seasonal features of time series and can fully explain seasonal autocorrelation and trends.^18^ Because the daily outpatient visits in this study exhibited seasonality, so the SARIMA model was used. The detailed modeling process was as the following four steps: First, determine the stationarity of time series. If not, the original time series should be transformed into a stationary one by the methods of common difference or seasonal difference. Second, examine the autocorrelation function(ACF) and partial autocorrelation function(PACF) to determine various parameters. Third, the lowest Bayesian information criterion (BIC) was used to examine goodness of fit and the residual sequence of an appropriately fitted model is white noise. Finally, optimal SARIMA model was adopted to fit the data for prediction.^19^

In this study, the SARIMA model was constructed with the data from January 1, 2023 to November 30, 2024.The optimal SARIMA model was used to predict the outpatient visits from December 1, 2024 to December 31, 2024. The effect of the model is evaluated by comparing the difference between the predicted value and the actual value. The mean absolute percentage error (MAPE) is used as the quality standard to evaluate the prediction accuracy of the model.^20^

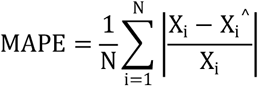

where X_i_ is the actual value at time i, X _i_ ^^^ is the predicted value at time i, and N is the number of data.

SPSS22.0. was used for the time series data processing and statistical modelling, and P < 0.05 was considered statistically significant.

## RESULTS

### Statistical description

Daily outpatient visits were collected from January 1, 2023 to December 31, 2024. In the past 24 months, a total of 1,720,875 cases have been reported. On the whole, the outpatient visits of the hospital showed an overall upward trend, with obvious seasonal fluctuation. Most notably, it exhibits clear trends and specific patterns throughout the week, known as weekly seasonality. Thus, we selected the daily time series for weeks 26, 52, and 78, based on the quartiles, and plotted a daily outpatient visits chart in the weekly time series. As shown in Figure 1, the number of outpatient visits on Monday and Saturday is significantly higher than that on the other five days of the week, the number of outpatient visits on Sunday is the lowest, while the number of outpatient visits on Wednesday is slightly higher than that on Thursday and Friday. The descriptive statistical analysis shows that the number of outpatient visits is a non-stationary time series with obvious periodic characteristics and weekly seasonal trends.

**Figure 1:**
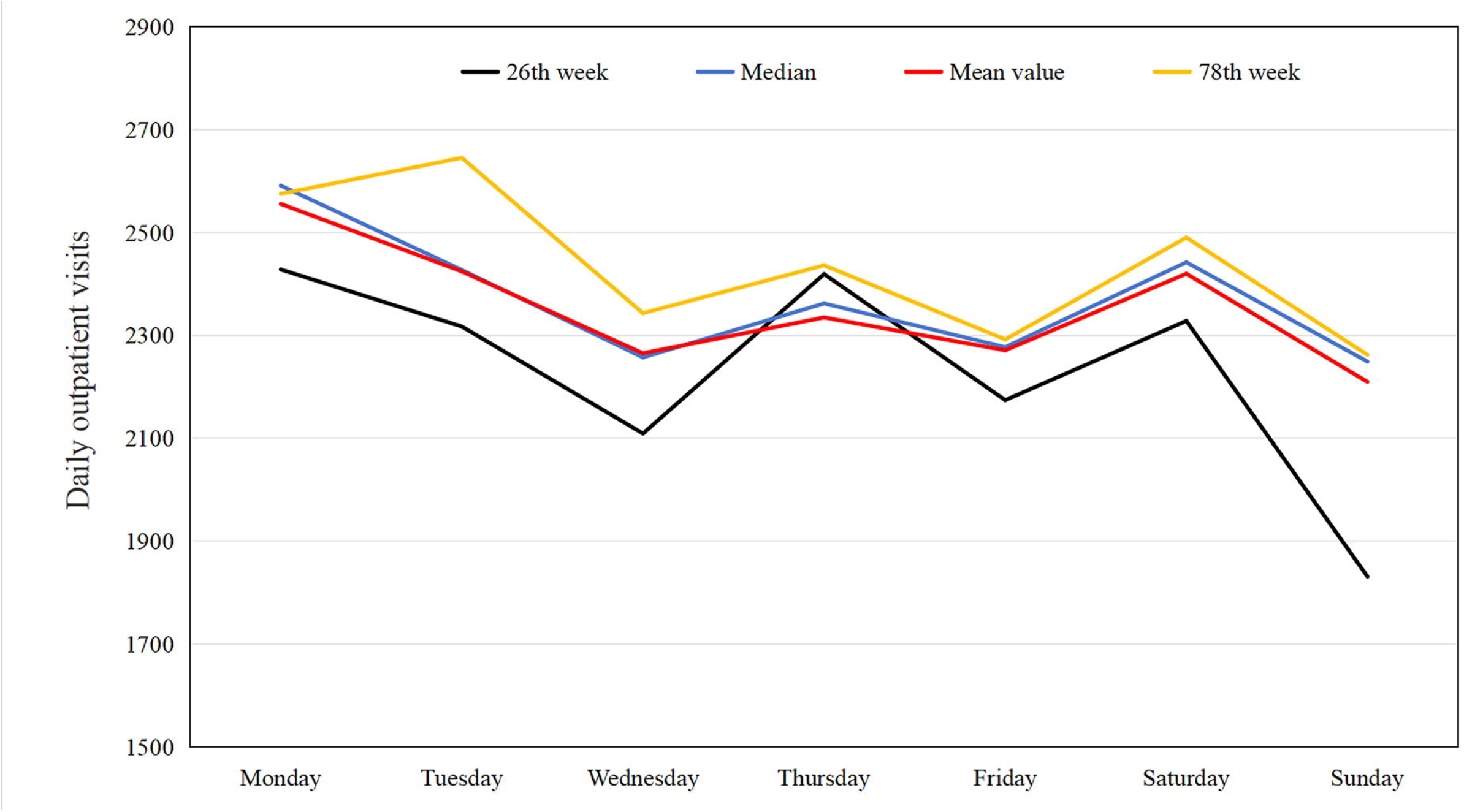
Daily outpatient visits chart in the weekly time series

### Model identification and diagnosis

Since the daily outpatient visits is a non-stationary time series with weekly seasonal trends, the SARIMA model is adopted. Using the raw training data from January 1, 2023 to November 30, 2024, trend difference (d = 1) and seasonal difference (D = 1) were completed. The original time series was transformed into a stationary one without any linear and seasonal trends. The auto-correlation Function (ACF) and partial correlation function (PACF) graphs were used to estimate the parameter ranges of p, P and q, Q as shown in Figure 2. After determining the model parameters, diagnosis is performed to show whether the parameters are statistically significant and to determine the optimal model. Several candidate SARIMA models were presented according to Table 1. On the basis of the results of the goodness-of-fit test statistics, SARIMA(1,1,0)(0,1,1)_7_ is the optimal model with the lowest BIC among the candidate models, the parameters of the SARIMA(1,1,0)(0,1,1)_7_ model are statistically significant according to the results of parameter estimation. This model also passed the Ljung-Box test (P > 0.05).

**Table1:**
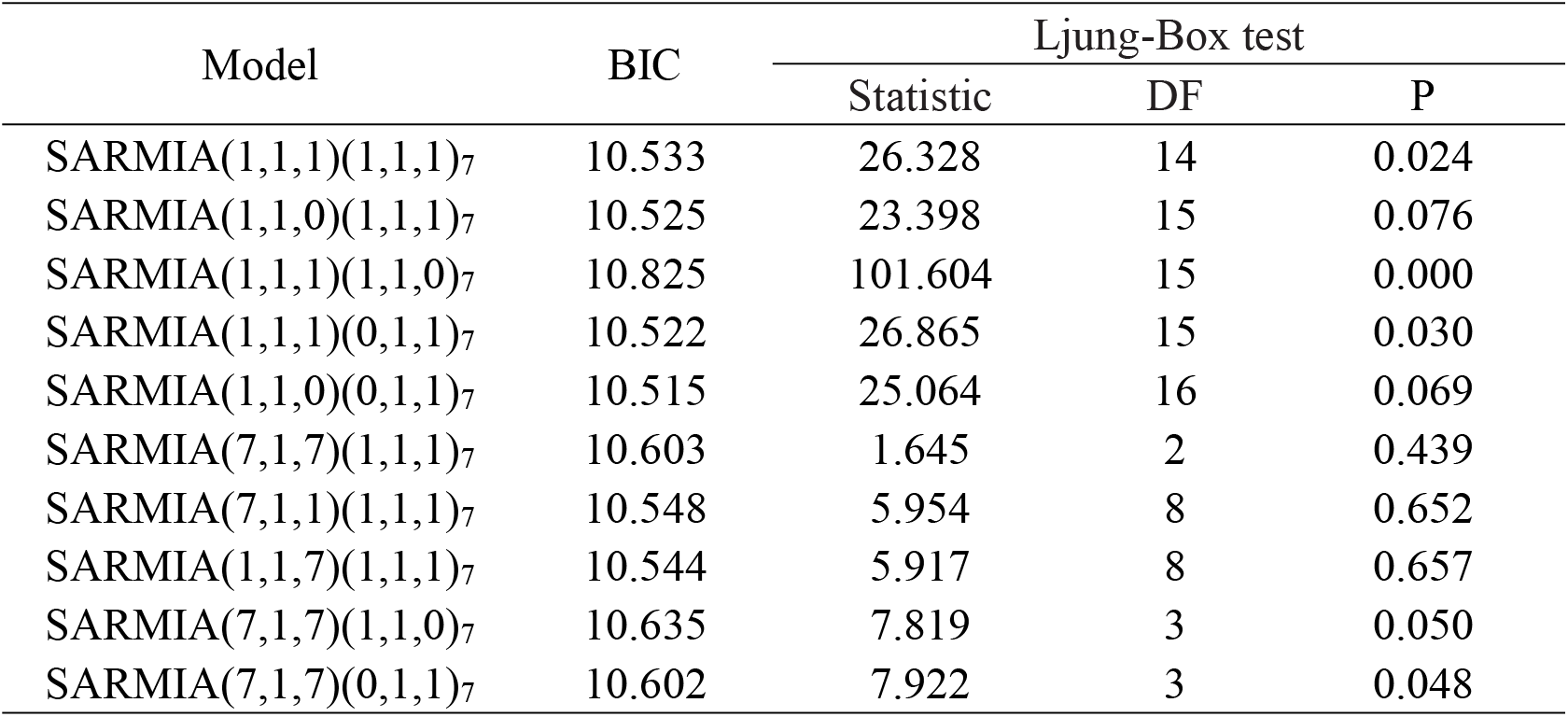
Goodness-of-fit of candidate models.

| Model | BIC | Ljung-Box test |  |  |
| --- | --- | --- | --- | --- |
|  |  | Statistic | DF | P |
| SARMIA(1,1,1)(1,1,1)<sub>7</sub> | 10.533 | 26.328 | 14 | 0.024 |
| SARMIA(1,1,0)(1,1,1)<sub>7</sub> | 10.525 | 23.398 | 15 | 0.076 |
| SARMIA(1,1,1)(1,1,0)<sub>7</sub> | 10.825 | 101.604 | 15 | 0.000 |
| SARMIA(1,1,1)(0,1,1)<sub>7</sub> | 10.522 | 26.865 | 15 | 0.030 |
| SARMIA(1,1,0)(0,1,1)<sub>7</sub> | 10.515 | 25.064 | 16 | 0.069 |
| SARMIA(7,1,7)(1,1,1)<sub>7</sub> | 10.603 | 1.645 | 2 | 0.439 |
| SARMIA(7,1,1)(1,1,1)<sub>7</sub> | 10.548 | 5.954 | 8 | 0.652 |
| SARMIA(1,1,7)(1,1,1)<sub>7</sub> | 10.544 | 5.917 | 8 | 0.657 |
| SARMIA(7,1,7)(1,1,0)<sub>7</sub> | 10.635 | 7.819 | 3 | 0.050 |
| SARMIA(7,1,7)(0,1,1)<sub>7</sub> | 10.602 | 7.922 | 3 | 0.048 |

**Figure 2:**
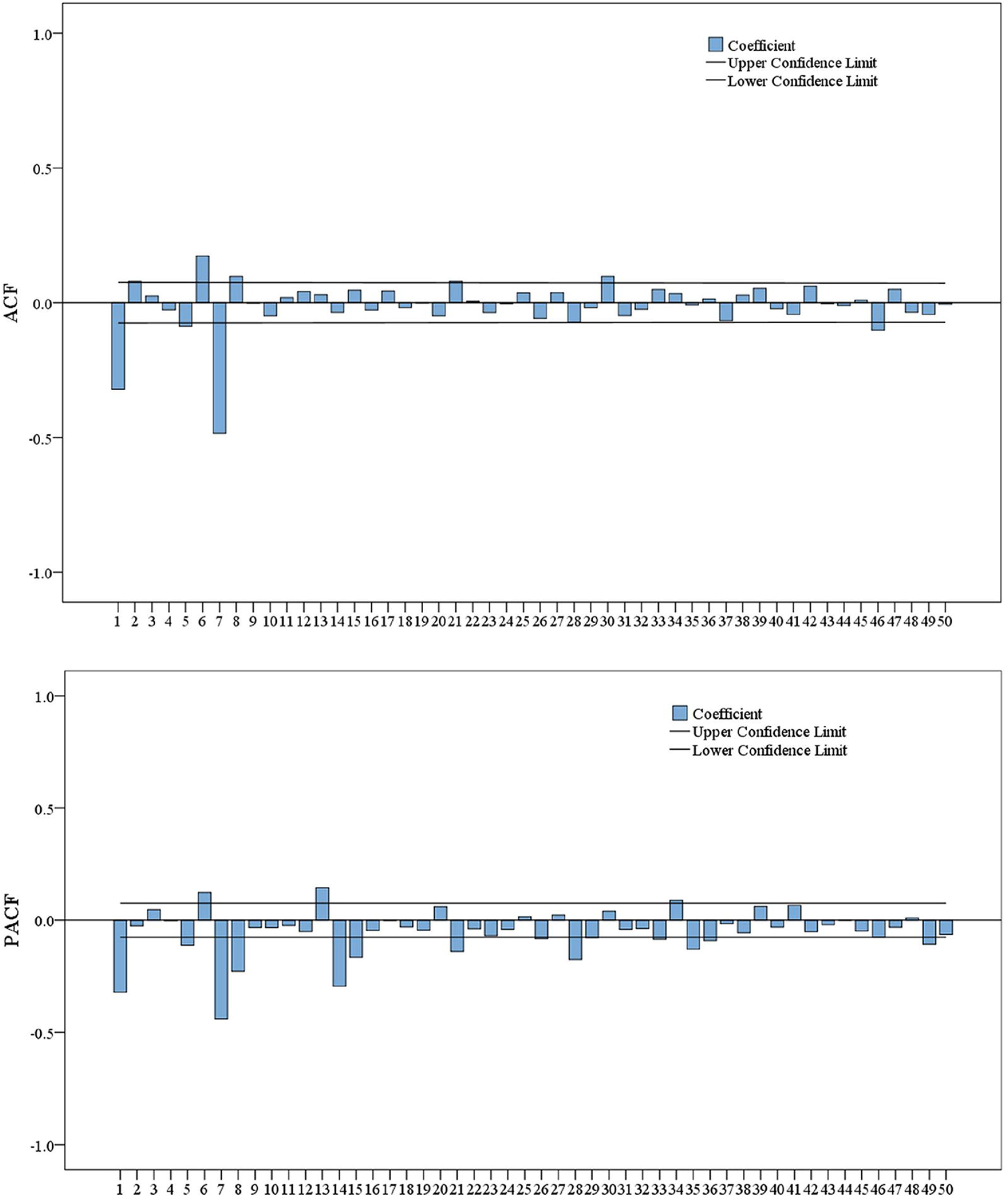
ACF and PACF graph of time series after once trend difference and seasonal difference

### Predictive evaluation

The SARIMA (1,1,0)(0,1,1)_7_ model fitting prediction graph is presented in Figure 3. The model exhibits excellent fit to the time series and was utilized to forecast the daily outpatient count for December 2024. The predicted values closely align with the actual values, as illustrated in Figure 4. The MAPE of the model is 0.073, indicating high prediction accuracy. As mentioned above, in our empirical research, the analysis results indicate that the model exhibits good predictive performance.

**Figure 3:**
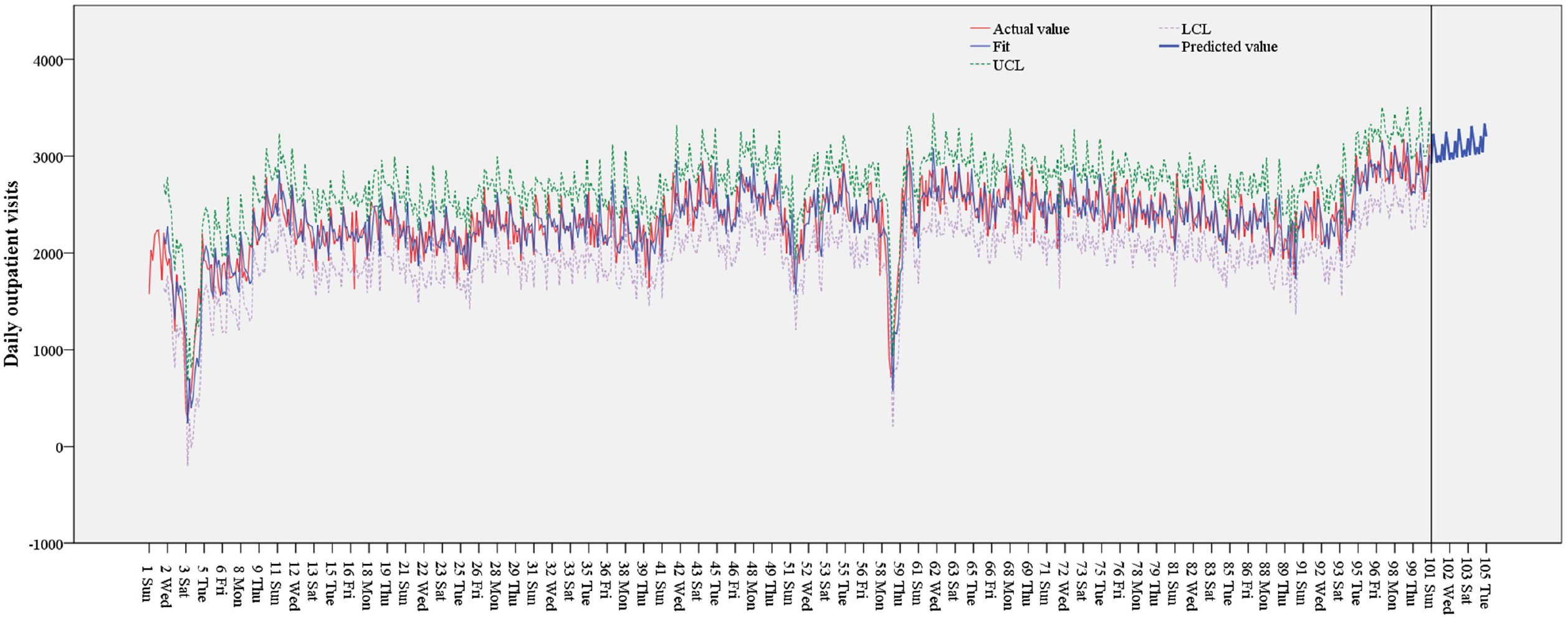
Fitting prediction graph of the SARIMA (1,1,0)(0,1,1)_7_ model.UCL, Upper confidence limit; LCL, lower confidence limit

**Figure 4:**
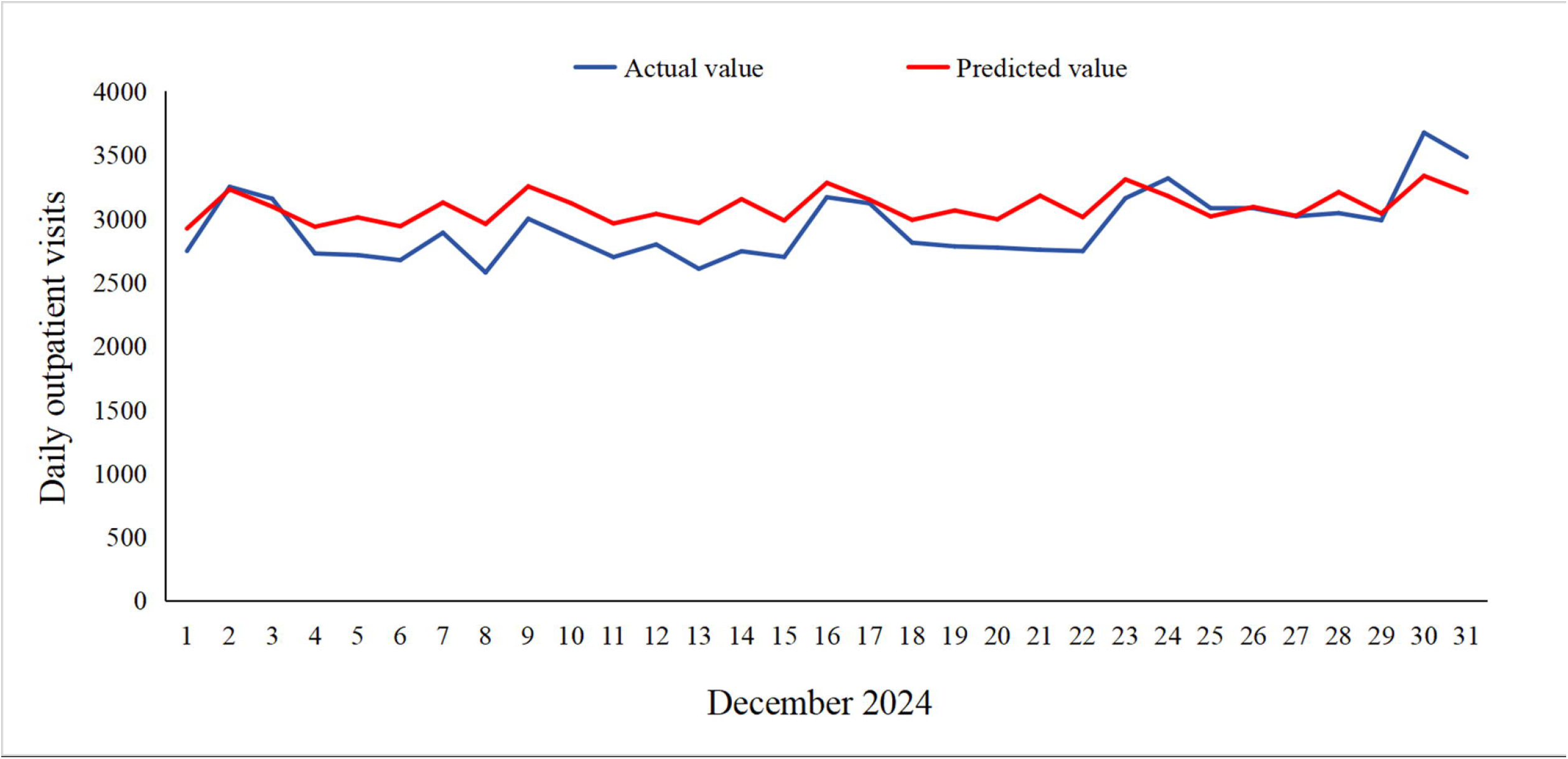
Comparison of actual and predicted daily Outpatient visits in December 2024

## DISCUSSION

In this study, the SARIMA model was employed to forecast hospital outpatient visits, with the objective of providing a scientific foundation for hospital management and optimizing medical resource allocation. The results of the SARIMA model showed promising predictive capabilities. It was able to capture the seasonal and trend components in the outpatient visit data. By analyzing and modeling outpatient data from January 1, 2023, to December 31, 2024, we obtained the predictive accuracy. This outcome not only offers robust support for hospital operational management but also serves as an empirical reference for applying time series analysis in healthcare settings. But in dynamic healthcare environments, accurate and timely forecasting is crucial. By incorporating short-term autocorrelation, periodicity, and the day-of-the-week effect into our model, we have developed a robust daily forecasting tool.

In terms of prediction accuracy, the SARIMA model demonstrated strong fitting capabilities. The MAPE of the predicted result is 0.073, indicating that the model’s daily predictions are very closely aligned with the actual outpatient visits. The error indicators fall within an acceptable range, suggesting a high degree of accuracy in the forecasting model’s performance. Furthermore, it could also fully utilize the information regarding the error term until it conforms to a white noise process. This indicates that the SARIMA model can effectively capture the characteristics and patterns of outpatient visits time series, enabling more precise future estimates. Many previous forecasting studies have relied on monthly or annual data.^21 22^ While medium and long term forecasts can achieve a good fitting effect, assist managers in making reasonable downstream decisions, and provide more buffer time to adapt to the cyclical schedule, they cannot offer adequate support for the precise allocation of medical resources at the operational level. Accurate short-term forecasts can offer theoretical support for managers to schedule human resources and prepare for emergencies, which is often more practical.^23 24^ In short-term forecasts, the model accurately reflects outpatient visits fluctuations. This is crucial for hospitals to efficiently schedule medical staff and deploy supplies, thereby enhancing service efficiency, reducing patient wait times, and improving overall patient experience. This information can be extremely useful for hospital administrators to plan staffing, allocate resources such as beds and medical equipment, and ensure smooth patient flow.

There are many factors that affect the number of outpatient visits in hospitals, such as personal experience, various environmental factors, and social factors. Changes in any of these factors may have an impact on the accuracy of the forecast for outpatient visits.However, obtaining this data can be relatively cumbersome, particularly for busy medical management personnel. Therefore, a quick and straightforward prediction tool becomes particularly crucial. The ARIMA model is relatively simple and can also be used to predict the number of outpatient visits without using covariates. In recent years, the ARIMA model has been widely used in the prediction of hospital. The time series analysis of 60-week daily visit data from the blood collection room of a large hospital in Chengdu shows that combined model of ARIMA and SES realize the short-to-medium-term prediction of the daily number of blood collections one week in advance.^25^ Chen, et al. utilized the ARIMA model to analyze and forecast the incidence rate and epidemic trend of gonorrhea in China.^26^ In Brazil, studies conducted by scholars have demonstrated that the SARIMA model predicts the number of dengue cases very effectively and reliably.^27^ Our research equally verifies the reliability of the SARIMA prediction model.

The prediction model results of this study not only exhibit high accuracy but also align with weekly trends, offering valuable guidance for daily operations. Specifically, it suggests that hospital managers may appropriately increase medical staff scheduling on Mondays and Saturdays. This increase should particularly focus on boosting the presence of renowned experts in outpatient clinics and enhancing the availability of diagnostic and treatment equipment. During Wednesdays, Fridays, and Sundays, when the number of outpatient patients decreases, health education efforts should be targeted at chronic disease patients, encouraging them to seek medical attention during these off-peak periods. This approach not only helps alleviate the pressure of high outpatient visits during peak hours but also shortens waiting times for patients. Consequently, it ensures adequate medical services during peak periods while minimizing unnecessary resource wastage during less busy times.

We acknowledge that our research has some limitations. It is important to note that the SARIMA model has its limitations. It assumes that the underlying patterns in the data remain relatively stable over time. In reality, holidays, unexpected events such as sudden disease outbreaks or changes in healthcare policies can significantly impact outpatient visits, and the model might not be able to account for these unforeseen circumstances fully. The factors mentioned earlier, including personal experiences, various environmental factors, and social factors, which influence outpatient visits, were not taken into account during the model fitting process. As a result, in the next step of research, these influencing factors should be incorporated into the study design to enhance prediction accuracy. Additionally, SARIMA is well-suited for short-term forecasting. As the prediction horizon widens, the data error rate tends to increase. Hence, SARIMA should be predominantly employed to forecast the most recent data.

## CONCLUSIONS

The SARIMA model can effectively simulate the trend of changes in hospital outpatient numbers over time series, providing a theoretical basis for short-term forecasting of hospital outpatient visits. By applying the time series model to the prediction of outpatient visits in hospitals, the variation rules of outpatient visits in different time periods can be analyzed with the help of the past outpatient visits data, and then the future outpatient visits can be scientifically estimated. This can not only help hospitals optimize internal resource allocation and improve operational efficiency, but also provide patients with a better and more efficient medical experience, which plays an important role in the stable operation of the entire medical system.

## Data Availability

All data relevant to the study are included in the article or uploaded as supplementary information.

## Acknowledgement

The authors thank Lei Zhang of the Hospital Information Department for his support in data collection.

## Ethical Approval

As this work focuses on statistical data of daily outpatient visits, direct data extraction from patients is not necessary. The need for ethical approval was therefore waived.

## Informed Consent

Not applicable.

## Provenance and peer review

Not commissioned; externally peer reviewed.

